# Interpretable Machine Learning based Detection of Coeliac Disease

**DOI:** 10.1101/2025.03.11.25323763

**Authors:** F. Jaeckle, R. Bryant, J. Denholm, J. Romero Diaz, B. Schreiber, V. Shenoy, D. Ekundayomi, S. C. Evans, M. J. Arends, E. Soilleux

## Abstract

**Background:** Coeliac disease, an autoimmune disorder affecting approximately 1% of the global population, is typically diagnosed on duodenal biopsy. However, inter-pathologist agreement on coeliac disease diagnosis is only 80%. Existing machine learning solutions designed to improve coeliac disease diagnosis often lack interpretability, which is essential for building trust and enabling widespread clinical adoption.

**Objective:** To develop an interpretable AI model segmenting key histological structures in H&E-stained duodenal biopsies, generating explainable segmentation masks, estimating intraepithelial lymphocyte (IEL)-to-enterocyte and villus-to-crypt ratios, and diagnosing coeliac disease.

**Design:** Semantic segmentation models were trained to identify villi, crypts, IELs, and enterocytes using 49 annotated 2048×2048 patches at 40x magnification. Subsequently, IEL-to-enterocyte and villus-to-crypt ratios were calculated from segmentation masks generated by the segmentation model from 172 whole slide images (WSIs), and a logistic regression model was trained to diagnose coeliac disease based on these ratios. Evaluation was performed on an independent test set of 613 WSIs from an independent medical institution.

**Results:** The villus-crypt segmentation model achieved mean PR-AUC of 80.5%, while the IEL-enterocyte model reached PR-AUC of 82%. The diagnostic model classified WSIs with 96% accuracy, 86% positive predictive value, and 98% negative predictive value on the independent test set.

**Conclusions:** Our interpretable AI models accurately segmented key histological structures and diagnosed coeliac disease in unseen WSIs, demonstrating strong generalization performance. These models provide pathologists with reliable IEL-to-enterocyte and villus-to-crypt ratio estimates, enhancing diagnostic accuracy. Interpretable AI solutions like ours are essential for fostering trust among healthcare professionals and patients, complementing existing black-box methodologies.

**What is already known on this topic:** Pathologist concordance in diagnosing coeliac disease from duodenal biopsies is consistently reported to be below 80%, highlighting diagnostic variability and the need for improved methods. Several recent studies have leveraged artificial intelligence (AI) to enhance coeliac disease diagnosis. However, most of these models operate as “black boxes,” offering limited interpretability and transparency. The lack of explainability in AI-driven diagnostic tools prevents widespread adoption by healthcare professionals and reduces patient trust.

**What this study adds:** This study presents an interpretable semantic segmentation algorithm capable of detecting the four key histological structures essential for diagnosing coeliac disease: crypts, villi, intraepithelial lymphocytes (IELs), and enterocytes. The model accurately estimates the IEL-to-enterocyte ratio and the villus-to-crypt ratio, the latter being an indicator of villous atrophy and crypt hyperplasia, thereby providing objective, reproducible metrics for diagnosis. The segmentation outputs allow for transparent, explainable decision-making, supporting pathologists in coeliac disease diagnosis with improved accuracy and confidence.

**How this study might a8ect research, practice or policy:** This study presents an AI model that automates the estimation of the IEL-to-enterocyte ratio—a labour-intensive task currently performed manually by pathologists in limited biopsy regions. By minimising diagnostic variability and alleviating time constraints for pathologists, the model provides an efficient and practical solution to streamline the diagnostic workflow. Tested on an independent dataset from a previously unseen source, the model demonstrates explainability and generalizability, enhancing trust and encouraging adoption in routine clinical practice. Furthermore, this approach could set a new standard for AI-assisted duodenal biopsy evaluation, paving the way for the development of interpretable AI tools in pathology to address the critical challenges of limited pathologist availability and diagnostic inconsistencies.

## I. Introduction

With the emergence of digital pathology, artificial intelligence (AI) has the potential to improve the accuracy and speed of diagnosis. Duodenal (small intestinal) biopsies in particular are well suited to develop novel AI algorithms for, as the percentage of malignant diagnoses (<1%) is small compared to other parts of the body.

The most common pathology found in the duodenum is coeliac disease, a chronic autoimmune disorder triggered by the ingestion of gluten, a protein found in barley, wheat and rye [1]. It is estimated to aFect approximately 1% of the global population, with significant regional variations [2,3]. The only available treatment is following a life-long gluten-free diet.

Coeliac disease can lead to variety gastrointestinal and systemic symptoms including vitamin deficiency, anaemia, weight loss, diarrhoea, fatigue, and infertility [4]. Furthermore, if left untreated it can significantly increase the risk of developing duodenal adenocarcinoma and lymphoma [4,5].

### Current diagnosis

Most guidelines suggest a serological test for patients with symptoms suggestive of coeliac disease. Unless the level of the tTG-IgA antibody count is exceeding 10 times the normal value and IgA EMA test positive in a second blood sample, a duodenal biopsy is recommended as the gold standard for coeliac disease diagnosis [6– 8]. Pathologists diagnose coeliac disease on microscopic sections of duodenal biopsies viewed under the microscope or as a scanned image, by identifying a number of abnormalities. These include an increased ratio of intraepithelial lymphocytes (IELs) to enterocytes in the villi, villous atrophy (significant blunting and reduction in size of the villi, sometimes to a flat appearance), and crypt hyperplasia (crypts becoming longer and thinner) [9]. However, a number of independent studies have showed the agreement between pathologists when diagnosing coeliac disease to be lower than 80% [9–19], underlining the unmet clinical need for a more accurate diagnostic solution.

### The Role of AI in Supporting Pathologists

Beyond the technical challenges, the development of AI tools for pathology must be aligned with the realities of clinical workflows and the human elements of diagnostic practice. Pathologists routinely face high workloads, and tasks such as quantifying intraepithelial lymphocytes or measuring villous atrophy are time-consuming, subjective, and prone to inter-observer variability. Rather than replacing pathologists, well-designed AI tools have the potential to support them, by automating repetitive tasks, offering consistent quantitative metrics, and highlighting diagnostically relevant regions, ultimately improving both efficiency and confidence in diagnosis. While this study focuses on developing and validating interpretable AI outputs, we view this as a foundational step toward building tools that could be integrated into real-world diagnostic workflows and accepted as practical aids by the pathology community.

### Other work in this area

Several recent studies have aimed to improve the diagnosis of coeliac disease on haematoxylin and eosin-(H&E)stained biopsies with AI. The majority used black-box classification algorithms that output a single diagnosis without providing any explanations or justifications which provides a significant barrier towards adoption: numerous studies on the acceptability of AI solutions in pathology emphasise that successful integration depends on making AI output “easily interpretable to clinicians and patients” [20–24]. Furthermore, most of studies did not test their models on an independent test set which would provide insights into its robustness and ability to handle common variability in image acquisition and staining procedures.

*Wei et al*. [25] trained a convolutional neural network (CNN) to classify individual 224×224 pixel patches as coeliac disease, normal, or non-specific duodenitis, achieving accuracies or 95.3%, 91.0%, and 89.2% respectively. However, they did not test their algorithm on an independent test set from a different source.

*Sali et al*. [26] trained a model to predict the Marsh-Oberhuber classification [27,28], an algorithm that classifies biopsies into different classes of coeliac disease severity. However, the study only included images from 34 patients.

*Seyd et al*. [29] developed a machine learning model to diagnose coeliac disease, environmental enteropathy (EE), and normal cases. However, the authors used different types of input images: EE images were digitized slides, whereas the coeliac disease and normal images were taken from a microscope at different resolutions. It is therefore possible that the model simply learned the image type rather than any pathological features.

*Koh et al*. [30] proposed a machine learning algorithm based on Steerable Pyramid Transform and entropy features to detect coeliac disease and classify its severity using Marsh scores. The most significant limitations of this study are the small size of the dataset and the lack of external validation on a dataset from a different source.

*Kowsari et al*. [31] proposed a hierarchical medical image classification approach using multiple deep learning models to classify small bowel enteropathies hierarchically. The model achieved high accuracy in classifying EE, coeliac disease, and normal controls, as well as coeliac disease severity. While promising, the study relied on a limited dataset from a single institution, which may not be representative of the broader patient population.

*Tabacchi et al*. [32] developed a Clinical Decision Support System for aiding in the diagnosis of coeliac disease. The system, named ITAMACDSS, employed a fuzzy classifier based on neural networks. The main weakness of the paper is that the test set is small (n=20) and comes from the same source as the training set.

*Denholm et al*. [33] trained a CNN using multiple-instance-learning (MIL) to diagnose whole slide images (WSIs) as normal or coeliac disease. The main limitation of this study is that it includes only clear cases of coeliac disease and normal duodenal mucosa and no ambiguous cases or images with different diagnoses which limit its real-life impact.

Jaeckle et al. [34] developed a CNN using MIL that diagnosed coeliac disease with accuracy comparable to that of expert pathologists. Their model was evaluated on a representative test set comprising consecutive duodenal biopsy samples. However, a key limitation of their approach is the limited interpretability of the model: its outputs could only be visualised through post hoc heatmaps, offering limited insights into the model’s decision-making process.

A few recent studies have attempted to use image segmentation techniques to diagnose coeliac disease in a more explainable manner.

*Gruver et al*. [35] used a customizing off-the-shelf AI software to segment villous epithelium, lamina propria, and crypts on H&E-stained biopsies and IELs and enterocytes on CD3 immunohistochemistry WSIs. The two main drawbacks are firstly that the cell detection is carried out on CD3-stained biopsies which are rarely used in routine reporting practice and secondly the algorithm is trained and tested on images from the same source, therefore not proving any generalisation performance.

*Griffin et al*. [36] created a CNN to identify cells, tissue, and artifact regions in biopsies of celiac disease and normal duodenal tissue. They extracted features from these regions and correlated them with Marsh scores. Weaknesses of their approach are the small size of the test set (n=28) and that the test set cases came from the same source as the training images.

*In this study we* developed an explainable model for the interpretable analysis of duodenal biopsies and the accurate diagnosis of coeliac disease. We summarise our main contributions as follows:

1. We trained segmentation models for duodenal biopsies that detect villi, crypts, enterocytes, and intraepithelial lymphocytes (IELs).
2. We used the model outputs to accurately predict the IEL-to-enterocyte ratio in duodenal biopsies, a key indicator for diagnosing coeliac disease.
3. We further use the model outputs to predict the villus-to-crypt ratio, that acts as a close approximation to the level of villous atrophy and crypt hyperplasia.
4. We trained a logistic regression classifier that takes as input the IEL-to-enterocyte and villus-to-crypt ratios and outputs an accurate coeliac disease diagnosis on a set of representative biopsies from a new source.

## II. Data

### Segmentation training dataset

We trained our segmentation models on 49 annotated patches (small sub-images taken from larger WSIs) at 40x magnification (approximately 0.25 microns per pixel), each sized 2024×2024 pixels, to ensure they were sufficiently large to be able to include multiple villi. The patches are taken from 43 unique WSIs (several patches were extracted from different parts of the same WSI) from five different medical centres scanned using four different scanners (see Table S1). Of the 49 patches in the training dataset, 27 have been extracted from WSIs with a previous normal diagnosis, 15 from coeliac disease WSIs, and 7 from WSIs with other diagnoses. We handpicked patches to ensure they included diverse regions of the biopsies and various cell types. Each patch includes annotations drawn by pathologists using the open source QuPath software [37] outlining the villous epithelium (150 unique annotations), crypt epithelium (759), villous enterocyte nuclei (13,853), crypt enterocyte nuclei (23,657), and IELs (4,368).

### Classification training dataset

We further trained a logistic regression model to classify coeliac disease versus normal diagnoses using the IEL-to-enterocyte and villus-to-crypt ratios as input features. We ran our segmentation models on 172 WSIs (77 from coeliac disease cases and 95 normal), sourced from four different institutions. The calculated ratios were then used to train the logistic regression model. This dataset only included clinical diagnoses and no further cellular annotations. A breakdown of hospital origins of these images is provided in Table S1.

### Classification test set

Finally, we used an independent test set from a source from which no training data had been included. The test set included 613 WSIs of which 88 had a coeliac disease (60% female, median age 47) and 525 a normal diagnosis (60% female, median age 62). The WSIs were scanned on a Philips IntelliSite Ultra Fast Scanner. The detailed patient demographic breakdown of the test set is provided in Table S2.

## III. Method

The overall pipeline for our proposed method (as highlighted in Figure 1) consists of three main stages: pre-processing, semantic segmentation and coeliac disease classification.

**Figure 1.**
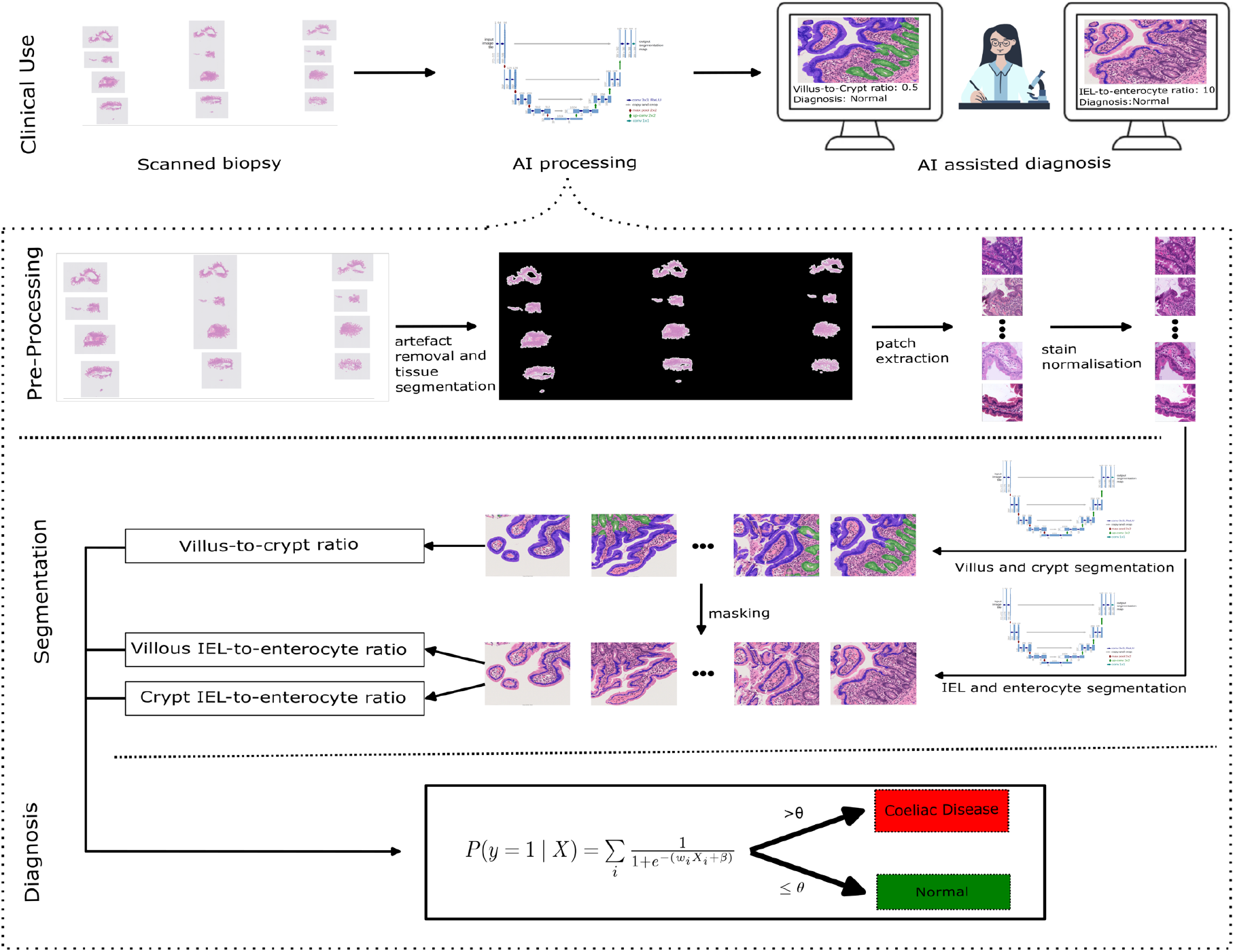
Model pipeline: The pipeline consists of the following steps: i) artefacts are removed, and the tissue is selected; ii) the tissue is divided into small patches. iii) stain normalisation is applied.; iv) villi and crypts are segmented using a U-Net model; v) IELs and enterocytes are segmented within the already segmented villi and crypts using another U-Net model; vi) the villus-to-crypt ratio and IEL-to-enterocyte ratios in both villi and crypts are computed; vii) a simple logistic classification model diagnoses the sample as either coeliac disease or normal. The outputs, including segmentation masks, the computed ratios, and the final diagnosis, are presented to a pathologist for review.

### III a. Pre-processing

We take inspiration from the work of Denholm et al. [33] to develop our pre-processing pipeline.

**Tissue Masking:** We used Schreiber’s method [38] to distinguish tissue areas from white background and unwanted artifacts and to generate tissue masks for the WSI.

**Patch Extraction:** Each WSI was divided into smaller patches at 40x magnification (the highest magnification available) of size 2024×2024 using QuPath.

**Stain Normalization:** To reduce variations caused by different staining and scanning procedures, we applied a stain normalization process using the Macenko method [39]. Stain normalisation has been shown by many studies to improve generalisation performance [40–42].

### III b. Cellular Segmentation

We trained two separate sets of semantic segmentation models: villus-crypt models and IEL-enterocyte models:

#### Villus-crypt model

we used the U-Net architecture [43] with a ResNet32 encoder [44] initialised with ImageNet pre-trained weights [45] and a final softmax layer applied on a pixel basis. We trained the models using 3-fold cross-validation for 70 epochs using the Adam optimiser with a cosine cycling learning rate scheduler (with a minimum and maximum learning rate of 1e-5 and 5e-5, respectively, updated at the end of every epoch) and using the focal loss [46]. We downsized each patch by a factor of two to 1024×1024 (at 20x magnification) and applied extensive data augmentation: random horizontal flips, rotations, and shifts, were applied to introduce spatial variability, while colour jittering (p=0.2) simulated changes in brightness [0,9,1,1], contrast [0,9,1,1], saturation [0.8,1.0], and hue [−0.5,0.5]. All code has been implemented in PyTorch (v1.13.1) [47].

#### IEL-enterocyte model

The IEL-enterocyte model focuses solely on the areas of the WSI that have already been segmented as either villi or crypts. During training we used the mask provided by the pathologist and during inference we used the output from the villus-crypt model to determine which parts of the image the models should focus on. We used the same model architecture and a similar set-up as before: we trained the models for 60 epochs and with a cycling learning rate scheduler where the minimum and maximum learning rates were 5e-5 and 5e-4, respectively. The most significant difference to the villus-crypt model was that here we split each patch into four smaller patches of size 1024×1024 at 40x magnification, as a form of regularization. As IELs and enterocytes are significantly smaller than villi and crypts, a higher magnification was needed.

### III c. Coeliac Disease Classification

Finally, we fit a simple logistic regression classifier that makes either a normal or coeliac disease diagnosis based on the masks produced by the two segmentation models. We first compute the villus-to-crypt area ratio, a surrogate metric for villous atrophy and crypt hyperplasia, by diving the number of pixels segmented as villi in the entire WSI by the number of pixels classified as crypt. Similarly, we compute the IEL-to-enterocyte ratio separately in the villi and the crypts. We then fit a simple logistic regression classifier that takes as input the three ratios and outputs a single diagnosis. Supplementary material D provides a more comprehensive analysis of the classification model.

## IV. Results

We first show how accurately we can predict the IEL-to-enterocyte ratio from segmentation masks. We then summarise the validation results of our segmentation before testing our segmentation and classification models on the independent test set.

### IV a. Predicting IEL-to-enterocyte ratios

As semantic segmentation masks do not specify instances of individual objects, but instead label each pixel as belonging to a type of object (e.g. an IEL), we can only estimate the number of instances of an object type (e.g. the number of IELs in a biopsy). We can predict the IEL-to-enterocyte ratio by dividing the number of pixels classified as IELs by the number of pixels classified as enterocytes in either crypts or villi whilst correcting for the different average cell sizes. As shown in Figure 2, Pearson correlation analysis showed that the ratio based on the total area covered by pathologists’ annotated IELs and enterocytes correlates very highly with the true IEL-to-enterocyte ratio derived from pathologists’ cell annotations (*r* = 0.92, *p* = 7.20−19). As pathologists do not count cells in the entire biopsy but rather in a few selected regions of interest, the mean absolute error of 0.016 ± 0.020 (which correspond to less than 2 IELs per 100 enterocytes) is not very significant. Predicting the IEL-to-enterocyte ratio based on the semantic segmentation masks is therefore justified assuming the mask is accurate.

**Figure 2.**
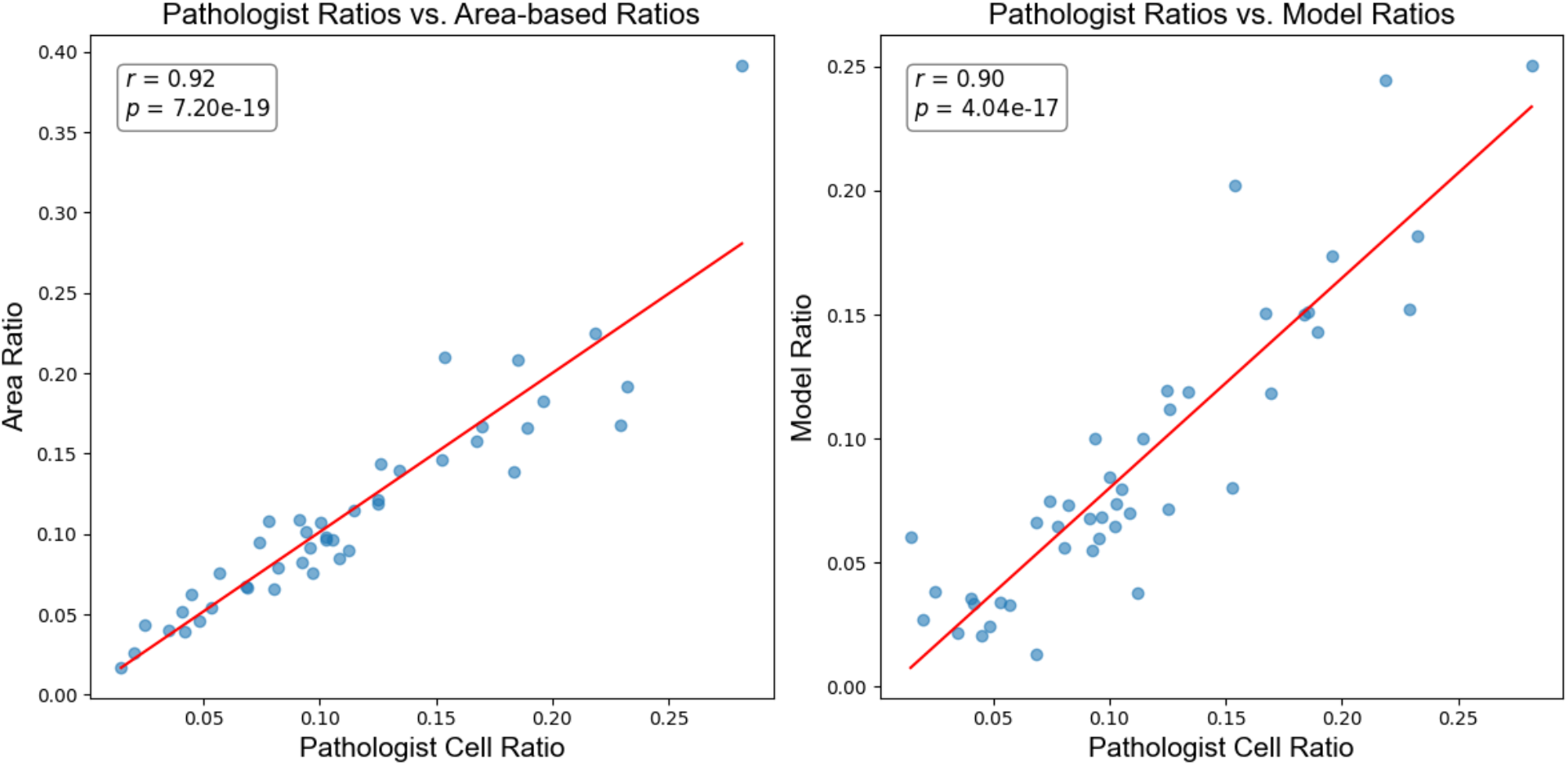
Comparisons of IEL-to-enterocyte ratio estimates. Left: The IEL-to-enterocyte ratio derived from pathologists’ cell annotations was compared to the ratio estimated based on the total area covered by pathologists’ annotated IELs and enterocytes. Pearson correlation analysis revealed a strong correlation with an r-value of 0.92 (p = 7.20 × 10^−19^). The mean absolute was 0.016 ± 0.02. **Right:** The IEL-to-enterocyte ratio from pathologists’ annotations was compared to the ratio estimated by the AI model on a validation dataset of 49 images. Pearson correlation analysis showed a strong correlation, with an r-value of 0.90 (p = 4.04 × 10^−17^). The mean absolute error was 0.028 ± 0.019.

### IV b. Validation of Semantic Segmentation Models

We trained and validated both the villus-crypt models and the IEL-enterocyte models using 3-fold cross-validation on the 49 annotated 2048×2048 patches. As summarised in Table S3, the villus-crypt models segmented villi with a mean accuracy of 90%, precision (positive predictive value) of 67% and recall (sensitivity) of 80%. The same models segment crypts with a mean accuracy of 90%, precision (positive predictive value) of 78%, and recall (sensitivity) of 71%.

The IEL-enterocyte models achieve mean accuracies of 91% and 99%, precisions (positive predictive values) of 85% and 65%, and recall (sensitivity) values of 85% and 67% for enterocytes and IELs, respectively, as summarised in Table S3. We include confusion matrices and some example patches in Figure 3, thereby further visualising the models’ validation performance.

**Figure 3.**
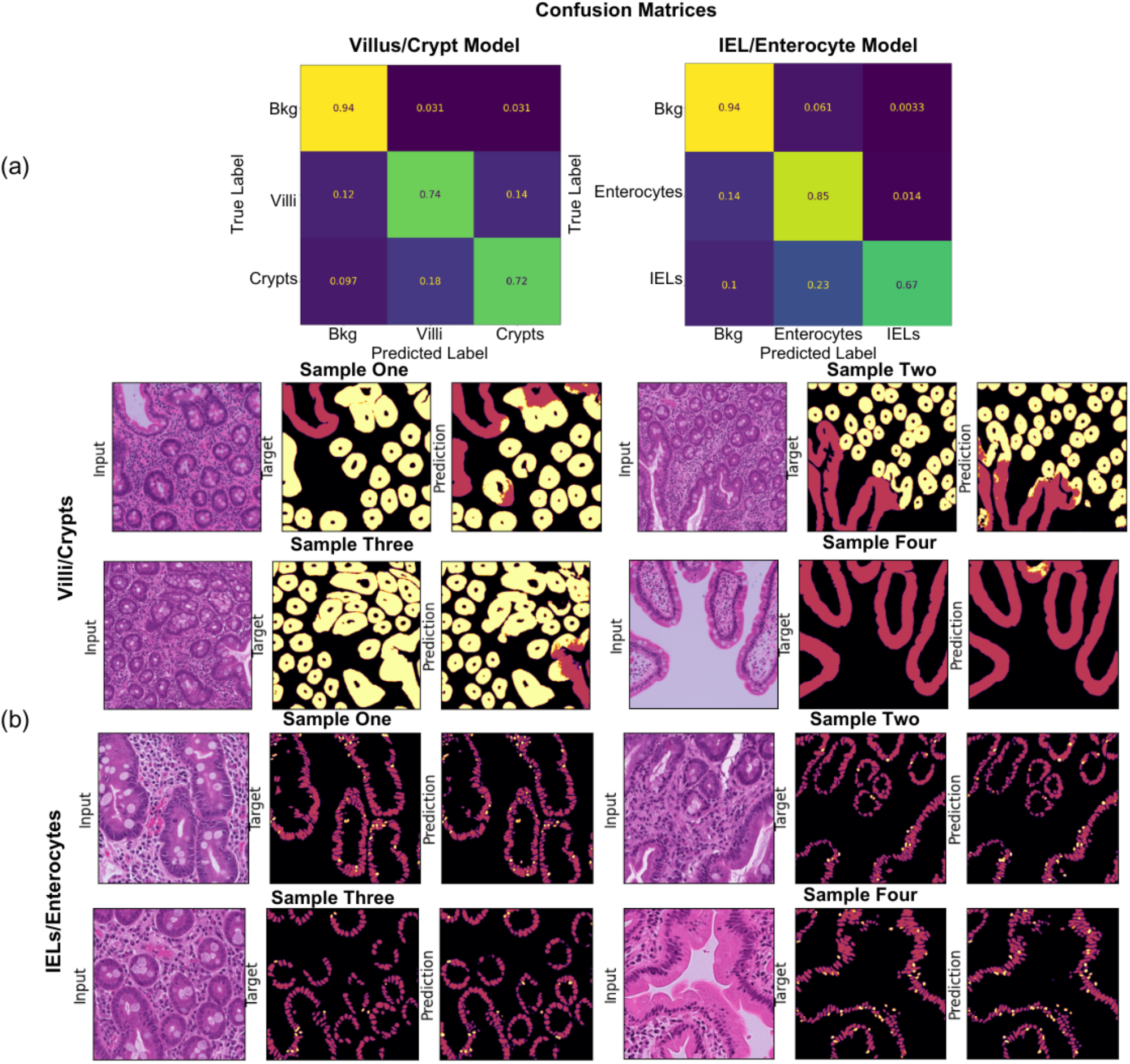
Validation Performance. a) Confusion matrix showing the mean validation performance of the two segmentation models on a pixel basis, normalised per true label (i.e., per row). For example, 72% of pixels annotated as crypts by pathologists were correctly segmented as crypts by the models. The most common misclassification was labelling IEL pixels as enterocyte pixels. b) Examples of validation images for each model set. Columns 1 and 4 display the raw image patches, columns 2 and 5 show pathologists’ annotations, and columns 3 and 6 show the corresponding model predictions.

Finally, we validated how accurately our segmentation models can estimate the IEL-to-enterocyte ratio, a key feature in the pathologist’s diagnosis of coeliac disease. As shown in Figure 2b, the IEL-to-enterocyte ratio estimated by the segmentation models only differs from the ratio calculated from the pathologists’ manual annotations by a mean absolute value of 0.028 (which correspond to 3 IELs per 100 enterocytes). Given that the threshold indicating coeliac disease used by pathologists varies between 0.2 and 0.4 (equivalent to 20 to 40 IELs per 100 enterocytes)[48–50], and the range of pathologist’s cell ratios across the patches spans from 0.02 to 0.28, we conclude that a mean error of 0.028 is acceptable and unlikely to be significant. The Pearson test further showed a strong correlation (*r* = 0.90, *p* = 7.20−19) between the pathologist’s and the model’s ratios. We further analyse the different IEL-to-enterocyte ratios in Appendix C.

### IV c. Test of the Coeliac Disease Classification Models

We also tested our coeliac disease classification pipeline that takes as input an entire WSI, then firstly carries out the pre-processing steps described in the methods section (background/artefact removal, patch extraction, stain normalisation), secondly runs the villus-crypt segmentation model, thirdly runs the IEL-enterocyte segmentation model, fourthly, computes the villus-to-crypt ratio as well as the IEL-to-enterocyte ratios in the villi and the crypts, and fifthly runs a classifier that takes as input these three ratios and outputs a diagnosis of either “normal” or “coeliac disease”.

We tested our segmentation models on a dataset comprising 613 WSIs from a previously unseen source to test the diagnostic accuracy and generalisation performance of our models. We visualise the output of the segmentation models on a few selected patches in Figure 4. As illustrated in Figure 5, the ratios we get for the normal and the coeliac disease populations are significantly different. The mean villus-to-crypt ratios were 0.91 ± 0.52 for the coeliac disease population and 2.83 ± 2.56 for the normal population (*p* = 7.0 × 10^−12^ using the independent samples t-Test), reflecting the significant level of villous blunting that can be observed in the majority of coeliac disease biopsies. Conversely, the IEL-to-enterocyte ratio in the villi is significantly higher (*p* = 2.4 × 10^−68^) for coeliac disease cases (14.44 ± 3.58) than it is for normal ones (7.81 ± 2.75), which matches clinical practice where pathologists use an increased IEL count in the villi as a key indication for coeliac disease. Interestingly, a statistically significant difference (*p* = 1.3*e*^−6^) is observed in the crypt IEL-to-enterocyte ratio, even though this metric is not typically considered by pathologists when diagnosing coeliac disease.

**Figure 4.**
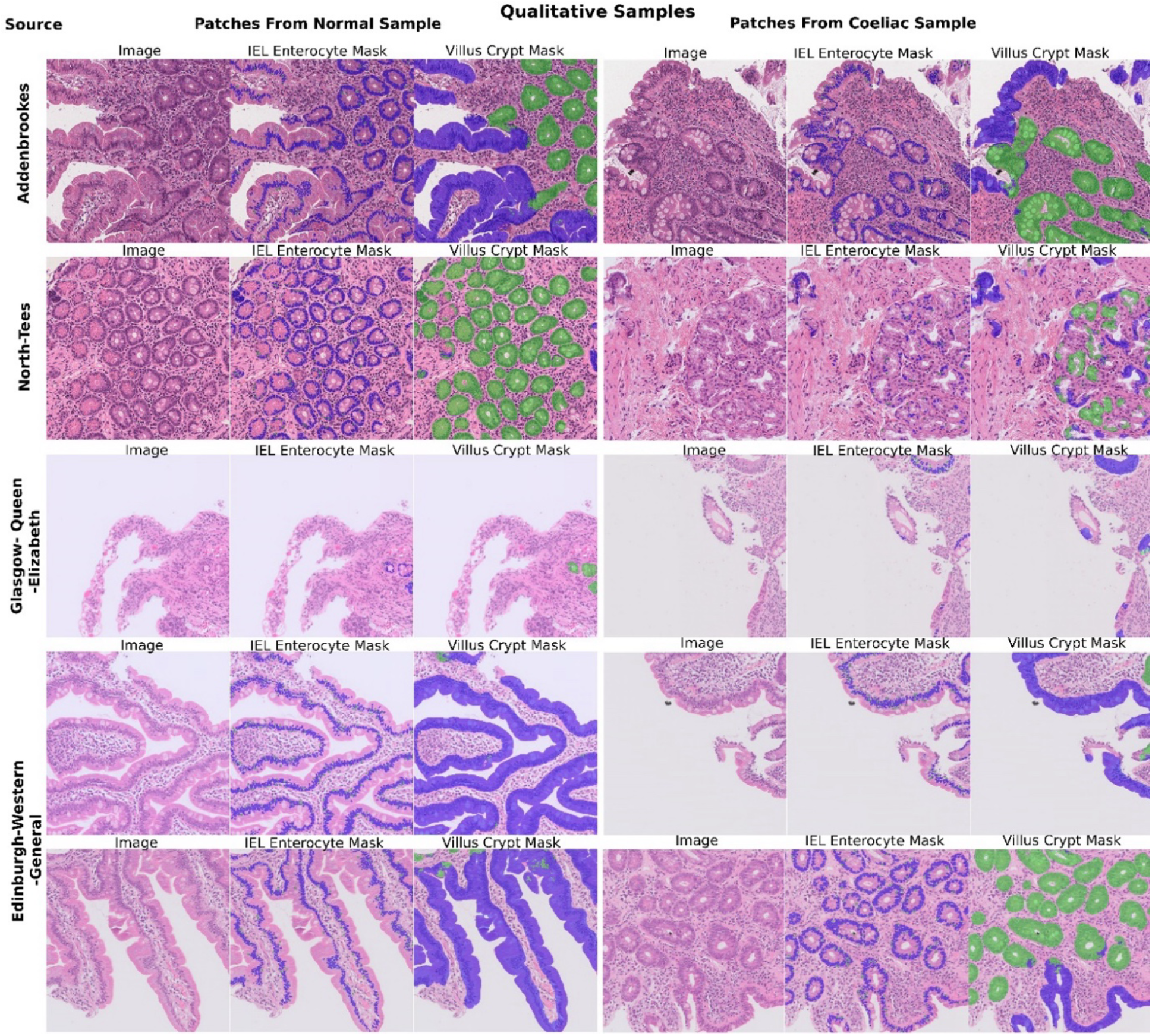
Sample visual outputs of segmentation models on test images: We showcase patches from 5 coeliac disease biopsies and 5 normal biopsies, collected from 4 diOerent hospitals. For each patch, three visualisations are provided: i) the original H&E-stained image, ii) the segmentation mask for IELs and enterocytes, and iii) the segmentation mask for villi and crypts. These outputs demonstrate the models’ robustness and eOectiveness across diverse sources and tissue characteristics.

Next, we evaluated the linear regression-based classifier on the same independent test set and achieved 96% accuracy, 86% positive predictive value (precision), and 98% negative predictive value, thus demonstrating how we can make an accurate explainable coeliac disease diagnosis. Importantly, we confirmed that our overall classification process for coeliac disease, based on semantic segmentation, followed by linear regression-based classification, achieved greater than 89% accuracy for all patient groups regardless of age and sex (Figure S6).

## V. Discussion

We have presented an accurate, explainable, and generalizable method for coeliac disease diagnosis in this work. We trained two sets of segmentation models to highlight villi, crypts, IELs, and enterocytes in duodenal biopsies. The segmentation masks can be reviewed by pathologists to develop trust in the model. We further showed how the model is able to estimate accurately the IEL-to-enterocyte ratio, one of the key characteristics on which pathologists base their coeliac disease diagnosis. We then demonstrated how the IEL-to-enterocyte and the villus-to-crypt ratios returned by the segmentation models are significantly different (p<0.00001) for the normal and coeliac disease populations. We subsequently trained a classifier that takes these ratios as input, and which gives a coeliac disease or normal diagnosis with over 94% accuracy. Unlike other previous works that aimed to train segmentation models for the diagnosis of coeliac disease, we tested our models on an independent test set, including 613 WSIs from a new hospital not previously used to train the models. We thus demonstrated very strong generalisability of our models which combined with the interpretable nature of our model outputs increase the likelihood of clinical adoption.

The work we presented marks an important step towards improving coeliac disease diagnosis for several key reasons. First, there is a global shortage of pathologists, resulting in long waiting times [51,52]. Second, studies consistently report high rates of disagreement among pathologists when diagnosing coeliac disease [9–19]. Third, most existing AI studies aimed at improving coeliac disease diagnosis function as “black boxes,” limiting their acceptance by healthcare professionals and patients due to a lack of transparency [25,26,29–33].

Our interpretable AI approach marks a significant first step toward software that could support faster, more accurate, and trustworthy coeliac disease diagnosis. Whereas pathologists typically estimate cell ratios from a few selected regions, our model performs a slide-wide, cell-by-cell analysis across the entirety of the biopsy, encompassing thousands or even tens of thousands of cells. To the best of our knowledge, this is the first explainable segmentation-based AI approach for coeliac disease diagnosis that has been evaluated on a large, independent test set from a separate clinical institution.

In a potential clinical workflow, a refined version of this software could be integrated into an image management system (IMS), where it would provide the pathologist with quantitative outputs including the IEL-to-enterocyte ratio, as well as a suggested diagnosis. These metrics would be displayed alongside the original biopsy image, allowing for rapid assessment and validation. To further aid diagnostic efficiency, the pathologist could request a set of top *n* patches with the highest IEL-to-enterocyte ratios, directing their attention to the most abnormal regions. This form of interaction maintains human oversight while leveraging the model’s analytical capabilities to support, rather than replace, clinical judgment.

Future work could extend the segmentation models to different types of cells, such as goblet cells, Paneth cells, or Brunner’s glands. Instance segmentation models may also return more accurate cell counts. The main limitation of this work is the lack of a perfect ground truth due to the imperfect nature of pathologists’ coeliac disease diagnosis. We hope that, in the future, our work can improve this situation, by providing more reproducible diagnosis of duodenal biopsies.

### V a. Limitations and Future Work

Tissue orientation during sectioning is a known source of diagnostic variability in histopathology, particularly in the assessment of villous and crypt architecture. In this study, we did not explicitly account for orientation or sectioning angle, which may lead to misrepresentation of crypt structures and impact the derived metrics (see for example the 3D model by Taavela et al. [53]). This limitation is especially evident in some poorly oriented slides (e.g., Figures 3, 4, and D.4). In future work, we aim to incorporate automated orientation assessment and filtering, or correction strategies, to ensure more reliable interpretation of morphometric features.

In this study, we used the ratio of segmented villi and crypt pixel counts as a surrogate metric for the villous height to crypt depth (VH:CD) ratio commonly assessed by pathologists. While this proxy does not replicate the exact methodology used in manual measurements, it offers a scalable and interpretable measure derived directly from the segmentation output. In future work, we plan to develop an algorithm that aims to replicate the clinical measurement process by identifying well-oriented villi and crypts within the segmentation masks and computing their average heights. This would enable a more direct and clinically meaningful comparison with pathologist-derived VH:CD ratios. Notably, despite the current simpler surrogate approach, our results in Figure 5 show a highly significant difference between normal and coeliac disease cases (*p* < 1e−^11^), highlighting the potential diagnostic relevance of the segmentation-based metric.

Beyond technical development, future work must also address how an improved interpretable AI tool would be deployed and evaluated in clinical settings. This includes prospective studies to assess its performance in real-world workflows, as well as qualitative studies to understand how pathologists perceive the tool’s utility, acceptability, and interpretability. Broader implementation questions, such as integration with image management systems, information technology (IT) infrastructure requirements, and energy consumption, are also key to ensuring that the solution is scalable and sustainable. Health economic evaluations will be important to assess the potential cost-effectiveness of automating labour-intensive tasks, reducing diagnostic variability, and potentially shortening time to diagnosis. These aspects are critical for the delivery of AI tools that are not only technically robust but also feasible and impactful in routine pathology practice.

## VI. Conclusion

This study presents an interpretable AI model that estimates key diagnostic features in coeliac disease, including the IEL-to-enterocyte and villus-to-crypt ratios (a surrogate metric for the villous height to crypt depth ratio) tasks that are currently time-consuming, subjective, and inconsistently performed by pathologists. By delivering a comprehensive, high-resolution analysis across entire biopsy slides, our approach provides objective and reproducible metrics that align closely with clinical criteria.

Tested on an independent dataset from a previously unseen institution, our model demonstrates strong generalisability and avoids the pitfalls of black-box systems by offering transparent, reviewable segmentation outputs. This allows pathologists to verify model predictions and focus their attention on diagnostically relevant regions, laying the foundation for clinical workflows that are both efficient and trustworthy.

Our interpretable AI approach marks a significant first step toward software that could support faster, more accurate, and consistent coeliac disease diagnosis. While additional work is required to evaluate the tool in live clinical settings and explore implementation at scale, this study provides a compelling proof-of-concept for how AI can meaningfully assist pathologists. In doing so, it helps to address the pressing challenges of limited workforce capacity, diagnostic variability, and increasing demand in modern histopathology.

## Data Availability

The datasets of WSIs analysed during this current study have not publicly available due to the large size of the WSIs and no patient agreements.

## Author’s Contribution

F.J., R.B., J.D., and J.R.D. developed the underlying methodology. E.S. conceived and set up the overall study. F.J. and R.B. drafted the manuscript. V.S. and D.E. created the cellular annotations for the training and validation dataset under the supervision of M.J.A.; J.D., B.S., V.S., S.C.E., and E.S. set up the annotation pipeline. All authors contributed to and had the opportunity to review and correct drafts of the manuscript.

## Acknowledgements

We would like to thank and acknowledge Graham Snudden for organisational support.

## Declaration of Interest

The authors declare the following financial interests/personal relationships which may be considered as potential competing interests: The following authors are shareholders in Lyzeum Ltd: F.J., J.D., M.J. A., and E.S.

## Funding

This work has been supported by Coeliac UK, Innovate UK (grant INOV03-19 to ES), NIHR (grant NIHR205502) and the Cambridge Centre for Data-Driven Discovery (C2D3).

## Ethics Approval

This study involves human participants and was approved. All slide scans (and accompanying fully anonymised patient data) were obtained with full ethical approval (IRAS: 162057; PI: Dr E Soilleux). Organisation: South Central – Oxford A (formally known as Oxfordshire Research Ethics Committee A).

## Notes

### Author Declarations

This study involves human participants and was approved. All slide scans (and accompanying fully anonymised patient data) were obtained with full ethical approval (IRAS: 162057; PI: Dr E Soilleux). Organisation: South Central - Oxford A (formally known as Oxfordshire Research Ethics Committee A).

### Summary of Updates

The Discussion was revised and a conclusion was added. No further results were added.

