## Supplementary for "Interpretable Machine Learning based Detection of Coeliac Disease"

#### Supplementary Material

Summary:

- Appendix A provides an in-depth analysis of the training, validation, and test data utilized.
- Appendix B presents detailed validation results of the segmentation model.
- Appendix C delves into the IEL-to-enterocyte ratio and further examines the pathologists' annotations used for training and validating the segmentation model.
- Appendix D offers a comprehensive analysis of the classification model, including the diagnostic threshold, confusion matrices for the validation and test sets, a detailed examination of the diagnostic model for various patient subgroups, an ablation study on the significance of different ratios, and illustrations of some outliers.
- Appendix G outlines the Patient and Public Involvement that guided this work..

[*] These authors contributed equally to this manuscript.

#### A. Dataset

We summarise the source of the training images as well as the scanner used to generate the WSIs in table S1. We then include a breakdown of the patient sub-populations of the test set in Table S2. The coeliac disease and normal diagnoses in made for the classification model training, validation, and test set were taken from the original clinical diagnosis made by NHS consultant pathologists. The annotations for the segmentation model training dataset were made by trainee pathologist and checked by a consultant pathologist.

| Source | Segmentation Model | Classification Model | Scanner |
|---|---|---|---|
| Addenbrookes | 15 | 60 | Leica Aperio AT2 |
| Glasgow-Queen-Elizabeth | 11 | 64 | Philips IntelliSite Ultra Fast Scanner |
| North Tees | 12 | 48 | Hamamatsu 210 |
| Heartlands | 4 | 0 | Roche Ventana iScan HT |
| Chaim Sheba Medical Center | 7 | 0 | Philips IntelliSite Ultra Fast Scanner |

**Table S1:** Breakdown of the training dataset used for training the segmentation and the classification models. We highlight both the source and the scanner. We note that the segmentation model was only trained on a 2048x2048 pixel patch from each WSI, whereas the classification model was trained on the entire image.

| | Subgroup | Coeliac (n = 88) | Normal (n=525) |
|---|---|---|---|
| Sex | Female | 53 | 313 |
| | Male | 35 | 212 |
| Age | median | 47 | 62 |
| | 0-9 | 12 | 15 |
| | 10-19 | 12 | 7 |
| | 20-29 | 5 | 18 |
| | 30-39 | 8 | 17 |
| | 40-49 | 13 | 68 |
| | 50-59 | 9 | 111 |
| | 60-69 | 19 | 109 |
| | 70-79 | 6 | 121 |
| | 80-89 | 4 | 57 |
| | 90-99 | 0 | 2 |

**Table S2:** Breakdown of the patient population for the test set based on sex and age.

#### B. Full Validation Results

We present the full validation results for the 3-cross-validation models for segmenting villi, crypt, enterocyte, and IELs. We report the accuracy, precision, recall, and PR AUC in table S3 below.

| Class | Fold | Accuracy | Precision (Positive Predicted Value) | Recall | PR AUC |
|---|---|---|---|---|---|
| **Villous Epithelium** | 0 | 0.94 | 0.56 | 0.91 | 0.85 |
| | 1 | 0.88 | 0.57 | 0.88 | 0.70 |
| | 2 | 0.88 | 0.88 | 0.60 | 0.87 |
| Mean | | 0.90 | 0.67 | 0.8 | 0.81 |
| Std | | 0.03 | 0.18 | 0.17 | 0.10 |
| **Crypt Epithelium** | 0 | 0.92 | 0.89 | 0.78 | 0.93 |
| | 1 | 0.90 | 0.85 | 0.58 | 0.76 |
| | 2 | 0.89 | 0.59 | 0.78 | 0.80 |
| Mean | | 0.90 | 0.78 | 0.71 | 0.80 |
| Std | | 0.02 | 0.16 | 0.12 | 0.04 |
| **Enterocyte** | 0 | 0.91 | 0.84 | 0.87 | 0.93 |
| | 1 | 0.90 | 0.87 | 0.82 | 0.93 |
| | 2 | 0.91 | 0.86 | 0.85 | 0.93 |
| Mean | | 0.91 | 0.85 | 0.85 | 0.93 |
| Std | | 0.00 | 0.01 | 0.02 | 0.00 |
| **Intraepithelial Lymphocytes (IELs)** | 0 | 0.99 | 0.66 | 0.68 | 0.72 |
| | 1 | 0.99 | 0.67 | 0.59 | 0.68 |
| | 2 | 0.99 | 0.63 | 0.74 | 0.73 |
| Mean | | 0.99 | 0.65 | 0.67 | 0.71 |
| Std | | 0.00 | 0.02 | 0.07 | 0.03 |

**Table S3.** Validation results from the Villus-Crypt and IEL-Enterocyte segmentation models.

#### C. IEL-to-enterocyte ratio estimation

We now focus on the IEL-to-enterocyte ratio in more detail. First, we explain how the pathologists annotated WSI patches; second, we analyse the sizes of the pathologist annotated cells; third we demonstrate how we can use the area of a patch covered by IELs and enterocytes to predict the cell count ratio; finally, we demonstrate how the model predictions correlate with the pathologist predictions.

##### C.1 Pathologist Annotations

Two different pathologists annotated all IELs and enterocytes found in 49 different 2048x2048 patches using QuPath. These annotations can be used to compute the IEL-to-enterocyte ratio in each of the patches. We analysed the distribution of cell sizes in Figure S1. We first plot the size of all IELs annotated by the pathologists. The mean size is 351 pixels (with a standard deviation of 134) which is roughly equivalent to 22 microns squared. The average villous enterocytes are $604 \pm 308$ and the enterocytes in the crypts are $691 \pm 346$.
We further plot the distribution of mean cell sizes per image in Figure S1. It shows a significant difference between average cell sizes between the different patches. We hypothesise that this is partly caused by the angle the tissue has been cut. To evaluate this we analyse the correlation of mean IEL and mean enterocyte size per image. We plot the patch-wise correlation in Figure S2. We conclude that the mean IEL and mean enterocyte size indeed heavily correlates, which could be an argument for the hypothesis that the cell sizes depend on the cutting angle. It further indicates that we will be able to estimate the cell ratio based on the cell area despite the irregularity between different patches.

Finally, we compare the average cell sizes for each annotator. The first pathologist annotated 15 slides and the mean cell sizes were as follows: $426 \pm 235$ for the villous enterocytes, $501 \pm 295$ for the crypt enterocytes, and $308 \pm 143$ for the IELs. The second pathologist, who annotated 34 patches clearly viewed the boundaries of the cells differently as they consistently created larger annotations. The mean cell sizes were as follows: $651 \pm 308$ for the villous enterocytes, $738 \pm 342$ for the crypt enterocytes, and $357 \pm 131$ for the IELs. This shows that different pathologists view the boundaries of these cells slightly differently. However, as our main is to predict the cell counts from the cell area, we simply require our model to be consistent.

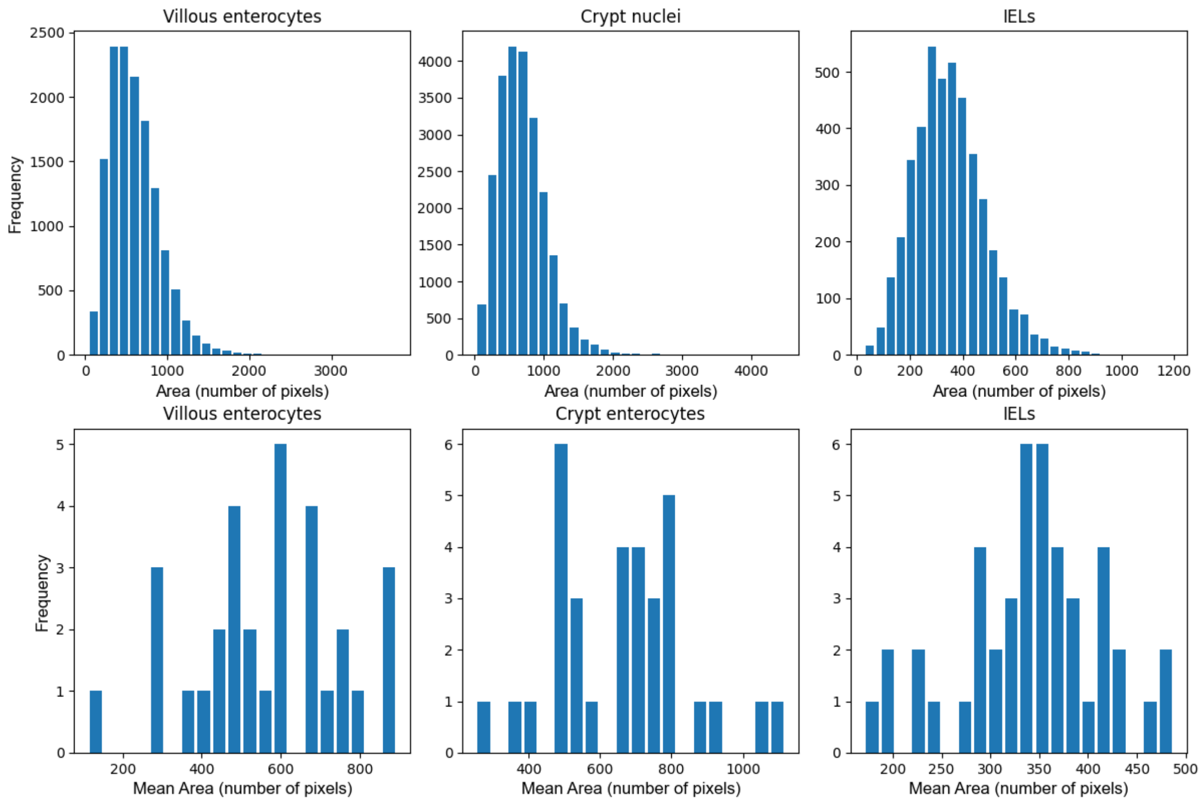


Figure S1. Top: we plot the distribution of different cell sizes as annotated by pathologists. Bottom: we plot the distribution of mean cell sizes per patch.

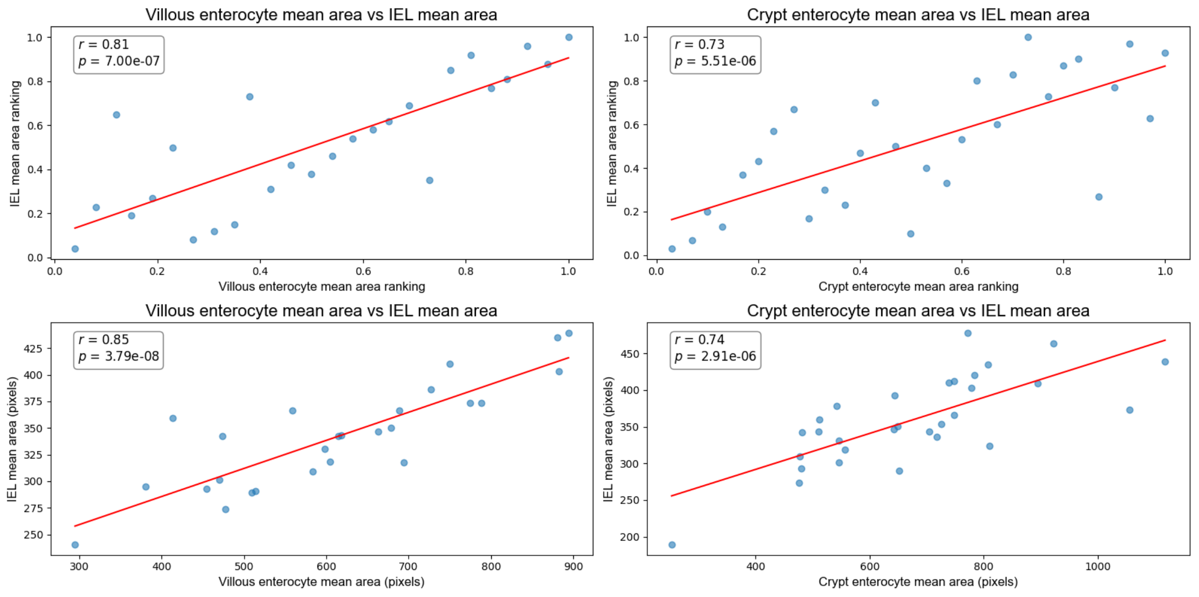


**Figure S2.** We highlight the correlation between the IEL and the villous enterocytes (left) and crypt enterocytes (right). As seen in the plots and as demonstrated by the Pearson correlation coefficient, there is a strong correlation between the IEL and the enterocyte sizes in each patch. We hypothesis that this is due to the way the slide is cut. We further note that this will allow us to estimate the cell count ratio from the cell area ratio, as the difference in cell sizes between the patches cancels out.

#### C.2 Area-based IEL-to-enterocyte ratio prediction

We now show how we can predict the IEL-to-enterocyte ratio from the ratio of the area of a patch covered by IELs, to the area covered by enterocytes. We predict the IEL-to-enterocyte ratio as follows

$$\text{IEL-to-enterocyte ratio} = \frac{A_{IEL}}{A_{V.E.} * \alpha_{V.E.} + A_{C.E.} * \alpha_{C.E.}},$$

where $A_{IEL}$ is the total area in number of pixels of a patch covered by IELs, $A_{V.E.}$ is the area covered by villous enterocytes, and $A_{C.E.}$ the area of crypt enterocytes. Furthermore, $\alpha_{V.E.}$ is a factor which is assigned to be the average IEL size divided by the average villous enterocyte size. Similarly, $\alpha_{C.E.}$ is a constant set to be the average IEL size divided by the average crypt enterocyte size. We note that to get a true validation performance without overfitting, we should not calculate $\alpha_{V.E.}$ and $\alpha_{C.E.}$ on the same patches that we calculate the IEL-to-enterocyte ratio on. We, therefore, split our validation dataset into two halves. We calculate the two constants on one half and use it to calculate the IEL-to-enterocyte ratio on the other half, and then we recompute the constants on the second half and use them to estimate the ratio on the first half. This way we get a more accurate validation performance. For the first fold we get $\alpha_{V.E.} = 0.606$ and $\alpha_{C.E.} = 0.501$ and for the second fold $\alpha_{V.E.} = 0.600$ and $\alpha_{C.E.} = 0.541$.

As shown in Figure S3 the IEL-to-enterocyte ratio predicted from the area correlates very highly with the true IEL-to-enterocyte ratio based on the number of cells annotated by the pathologists. The Pearson coefficient of $r = 0.92$ further highlights this. This result clearly motivates the use of the cell areas to predict the cell ratios.

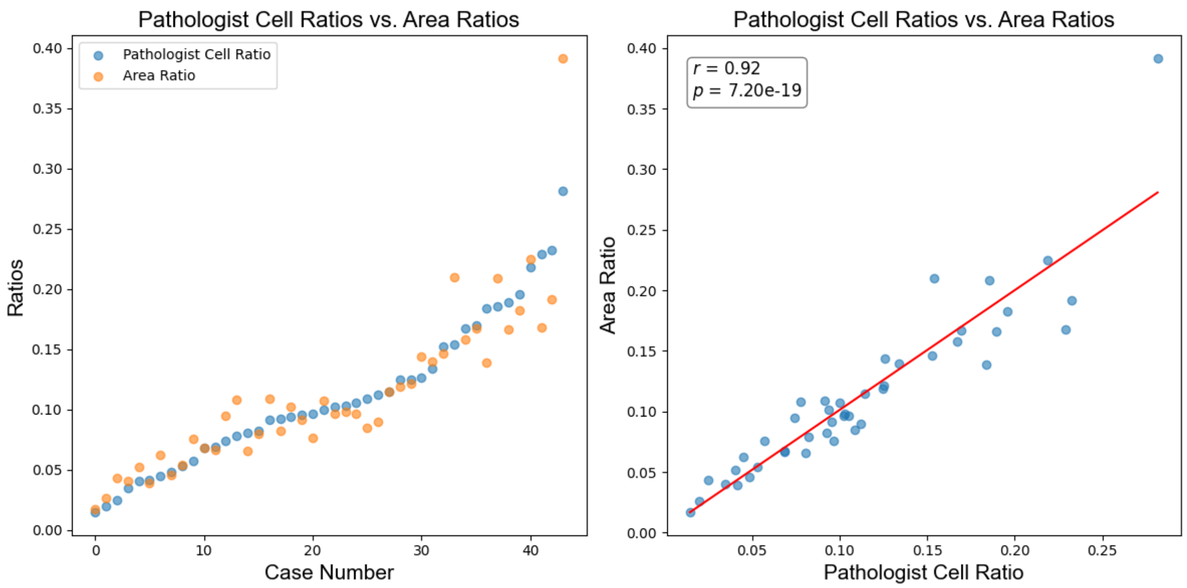


**Figure S3.** We plot the correlation between the IEL-to-enterocyte ratio calculated from the number of IEL and enterocyte ratios made by the pathologist in each patch, with the ratio computed based on the area covered by the annotations of each type of cell. The ratio estimated using the cell area correlates very highly with the ratio based on the annotation counts.

#### C.2 Model IEL-to-enterocyte ratio prediction

We now compute the IEL-to-enterocyte ratio based on the outputs of the segmentation model. We use the same two folds and the same constants $\alpha_{V.E.}$ and $\alpha_{C.E.}$ as in the previous section. We show the correlation between the model ratio and the pathologist ratio in Figure S4. We further computed a Pearson correlation coefficient of 0.9, indicating that the model very accurately predicts the IEL-to-enterocyte ratio.

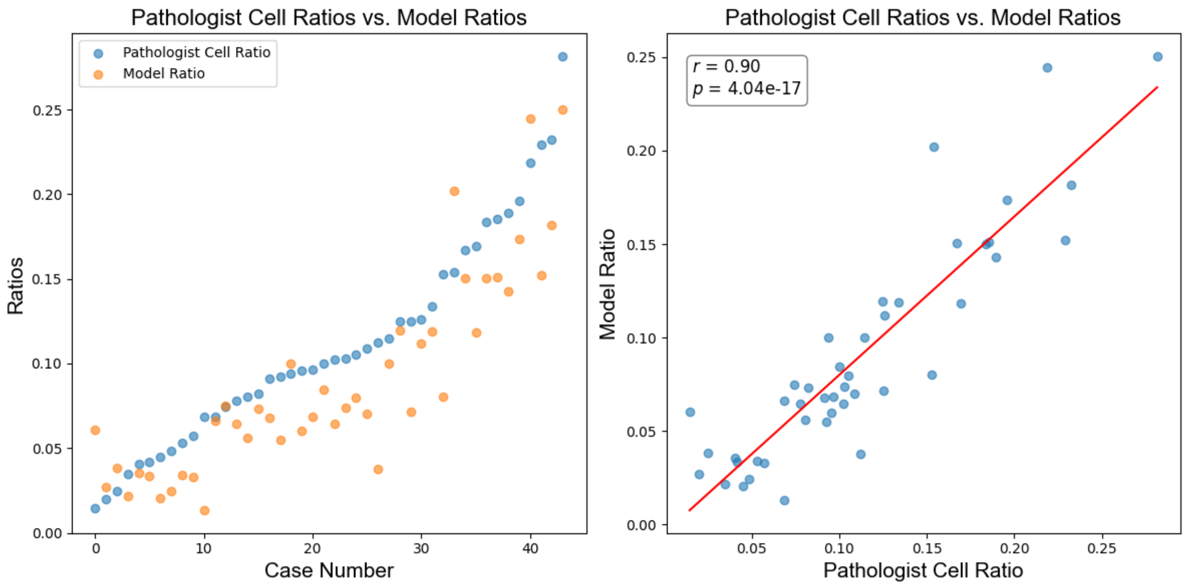


**Figure S4.** We plot the correlation between the IEL-to-enterocyte ratio calculated from the number of IEL and enterocyte ratios made by the pathologist in each patch, with the ratio computed based on the outputs of the machine learning model. The ratio estimated using the model correlates very highly with the ratio based on the annotation counts.

### D. Classification Model

#### D.1 Thresholding

The logistic classifier takes as input the three ratios outputs a number between 0 and 1, were 0 corresponds to a normal biopsy and 1 to a slide that shows strong signs of coeliac disease. We use a threshold $\theta$ to determine which raw classifier outputs lead to a normal diagnosis and which to a coeliac disease diagnosis. The lower the threshold, the higher the NPV and the lower the PPV as we return more coeliac disease diagnoses. Conversely, a higher threshold leads to fewer coeliac disease diagnoses and consequently a higher PPV and lower NPV. Depending on the use case, our model can thus be calibrated without needing any further training. We set our threshold to the value ($\theta = 0.7$) that optimises the sum of the PPV and NPV on the validation dataset. Figure S5 plots the accuracy, PPV, NPV, and PPV-NPV-average on the validation dataset. We further plot the same metrics for different thresholds on the test dataset. We note that this latter is for illustrating purposes only as in most cases one cannot change or optimise the threshold on the test set as it would then no longer be a true independent test performance. However, we include the plot as there may be use cases where a model can be fine-tuned when used in a new setting.

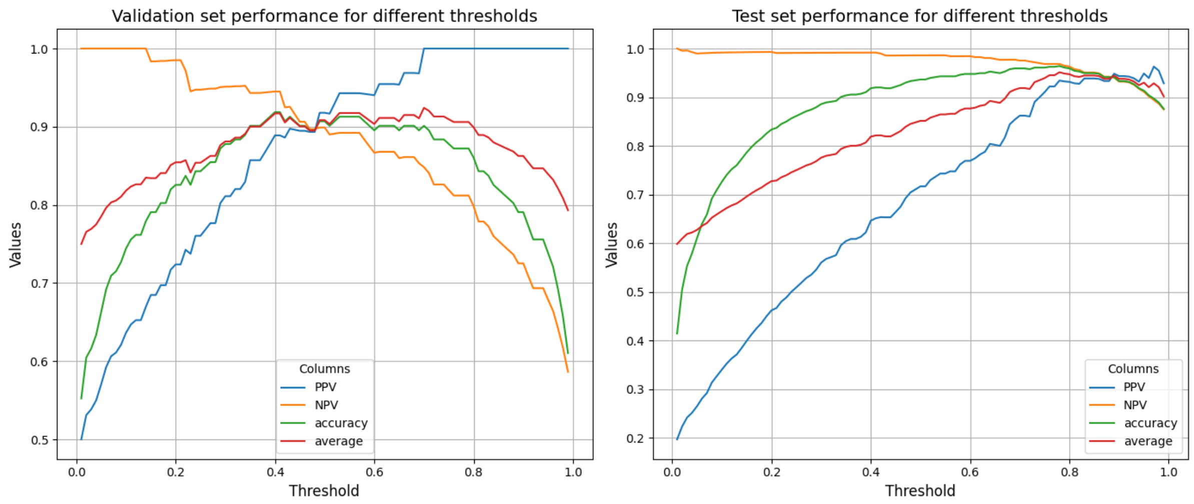


**Figure S5.** Threshold plots on the validation (left) and the test set (right). By adapting the diagnostic threshold, we can increase either PPV, or NPV depending on the application without needing to retrain the model.

#### D.2 Confusion Matrices

We plot confusion matrices for the validation and test set in Figure S4, highlighting the accuracy and the misclassifications.

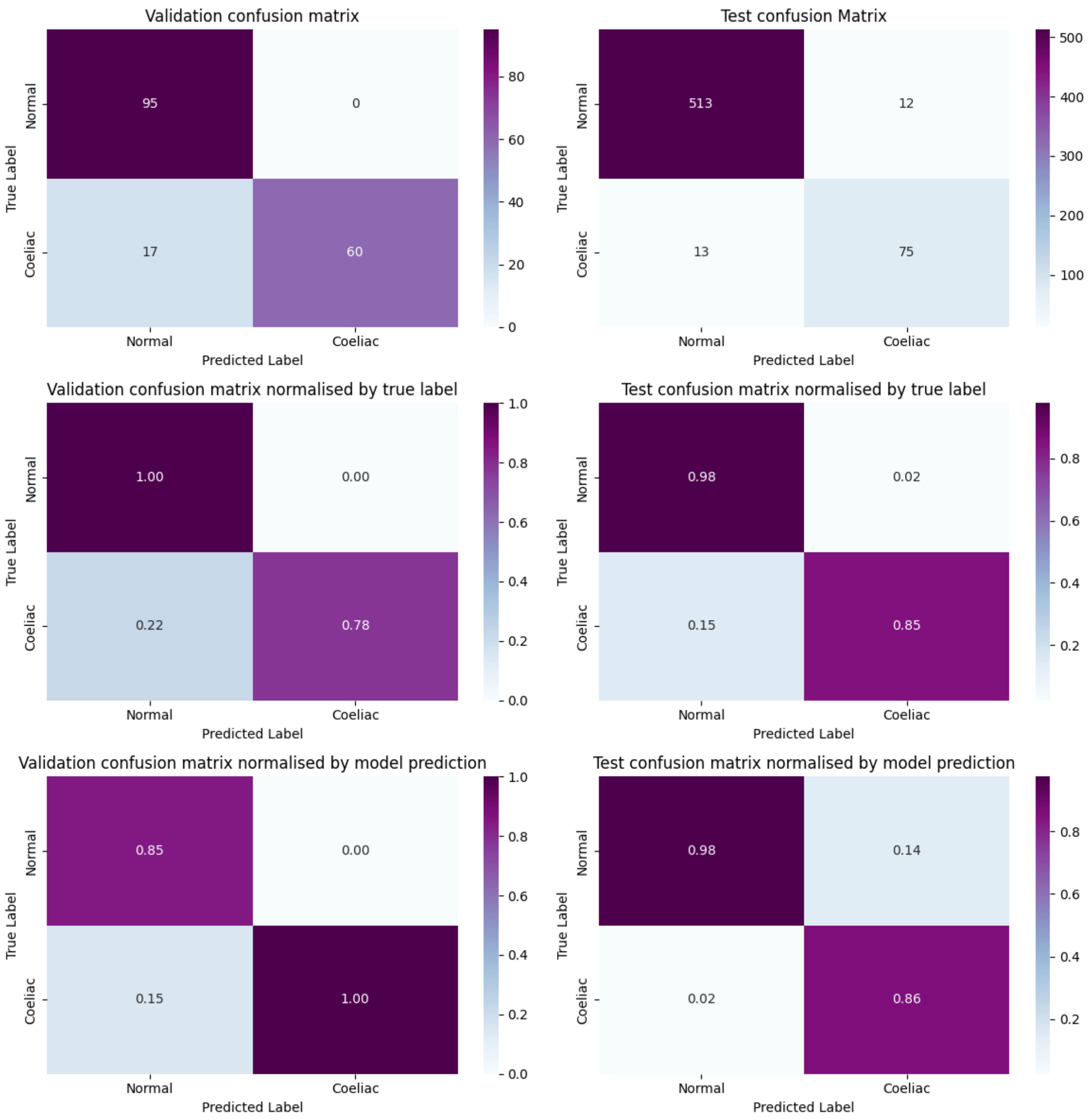


**Figure S4.** Confusion matrices for the coeliac disease diagnosis made by the logistic classifier on the validation set (left) and the test dataset (right). The first row includes raw confusion matrices, the second row contains confusion matrices normalised by the true label (the diagnosis made by the pathologist), and the third row contains confusion matrices normalised by the model predictions.

#### D.3 Performance by subgroup and ablation study of ratio importance

In order to ensure that our model works for all patient subgroups, we analyse the accuracy, PPV, and NPV on patients of differences ages and sexes. Our model performs well with over 89% accuracy for all subgroups. It performs equally well for both male and female patients. It is more accurate for patients above the age of 44 than for those under 44. The PPV for patients above the age of 75 is lower than for the remaining patients, which may be partially caused by the fact that very few patients above the age of 75 get diagnosed with coeliac disease.

We further show an ablation study in Figure S6. We compare the main model with three smaller models that take as input only one of the three ratios. We summarise that the model that takes as input all three models is the strongest. The villus-to-crypt ratio model is the strongest out of the remaining models, with the crypt IEL-to-enterocyte model being significantly less accurate. This matches both the distribution of ratios plotted in Figure 5 and also the conventional wisdom that villus atrophy and the IEL-to-enterocyte ratio in the villi are the key diagnostic markers, where pathologist don't regularly count IELs in crypts.

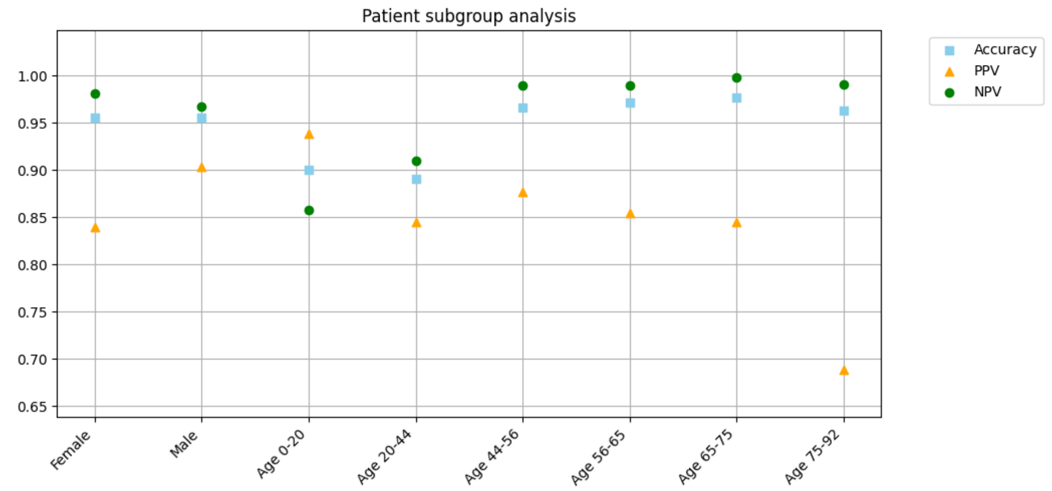


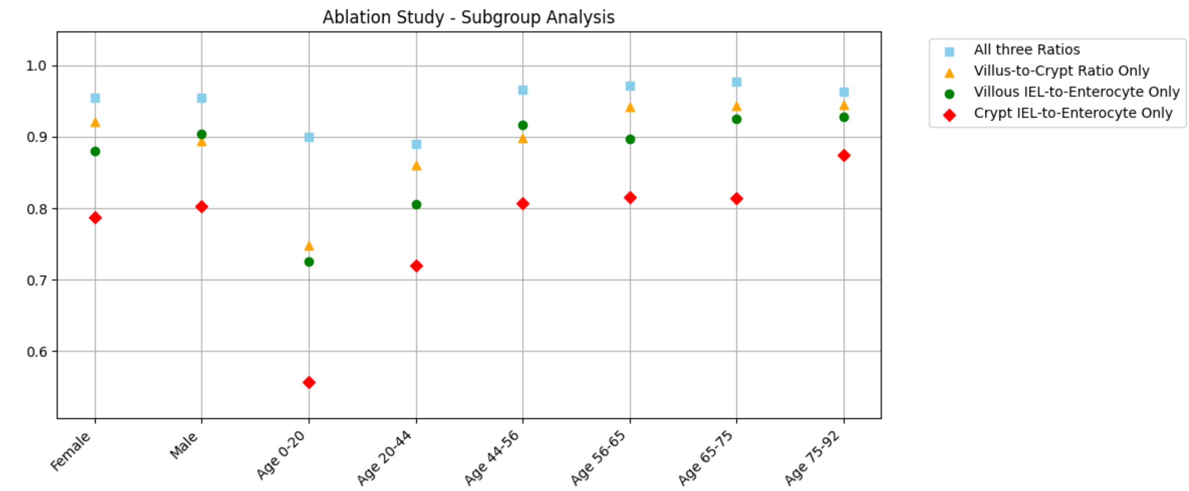

**Figure S6.** Top: we highlight the performances of our diagnostic classification model on various patient subgroups on the independent dataset. We plot the model's accuracy, PPV, and NPV, for patients of different age groups and different sex. Bottom: we show the main model described in the paper that takes as input three ratios (Villus-to-Crypt, Villous IEL-to-enterocyte, and Crypt IEL-to-enterocyte) and compare it to separate models that i) only take the villus-to-crypt ratio as input (yellow), ii) only take the IEL-to-enterocyte ratio in the villi as input (green), and iii) only take the IEL-to-enterocyte in the crypts as input.

#### D.4 Outliers

As can be seen in Figure 5, there were three outliers in the test set that had a very high villus-to-crypt ratio. All of these were normal cases, so this did not lead to misclassifications. As can be seen in Figure S7, the algorithm did segment the villi and crypts correctly and the high villus-to-crypt ratio is calculated correctly.

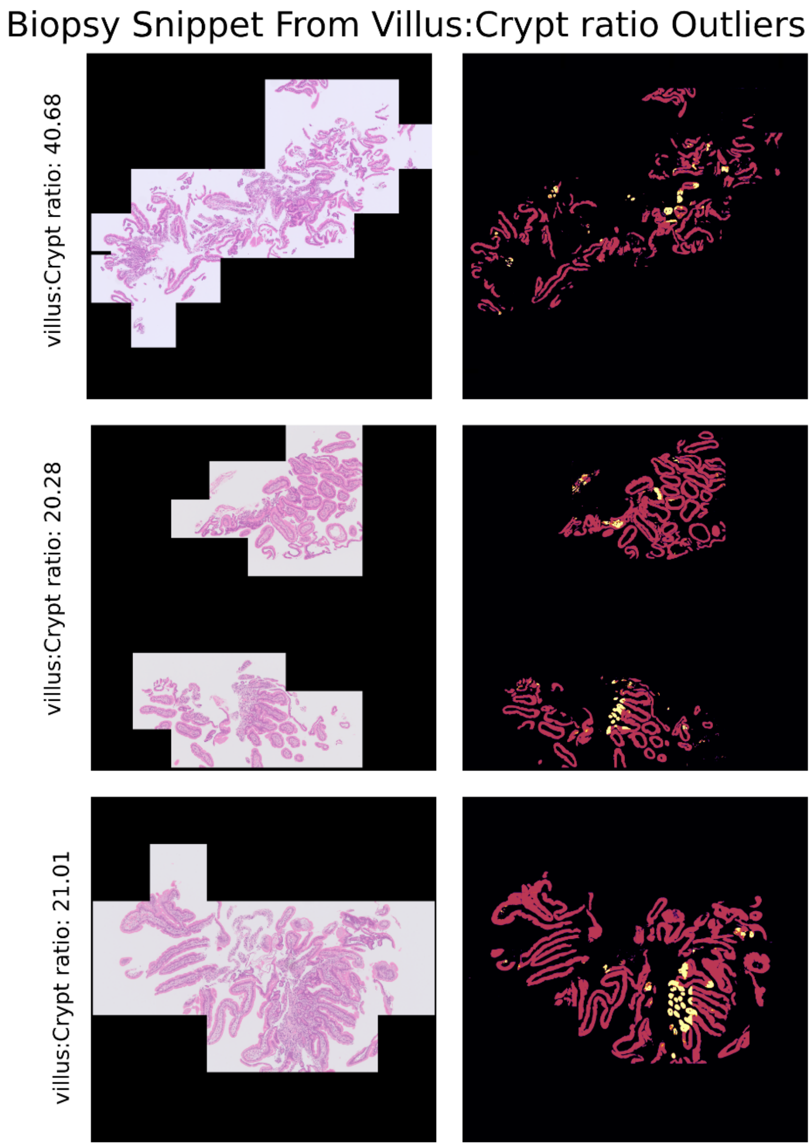


**Figure S7.** This figure shows high resolution samples from the outliers identified in Figure 5. The complete masks generated for sections of the outlier samples. These results demonstrate that the observed anomaly is not due to an error in the masking process; rather, the slides genuinely exhibit an unusually high number of villi. This could be attributed to the angle at which the biopsies were cut or the orientation of the tissue on the slide. Nevertheless, the critical takeaway is that our algorithm is performing correctly, and the observed outliers do not indicate a flaw in the method, which is a key success for this project

#### E. Patient & Public Involvement

We have run patient and public involvement workshops with Coeliac UK, the UK's leading charity for people suffering from coeliac disease. A survey of 198 members of the charity, 92% of whom had been diagnosed with coeliac disease, revealed that 70% were happy for a health care practitioner to rely on AI when diagnosing coeliac, with only 5% against it. Furthermore, 2 out of 3 would either strongly or very strongly trust a diagnosis of coeliac disease made using AI. However, the concern that came up most frequently once a group of 30 members had heard a presentation about existing AI tools, was the black-box nature of a lot of AI algorithms. This research aims to address this concern by developing more explainable AI solutions for the diagnosis of coeliac disease.
